# Effects of age and lesion characteristics on chronic outcomes after childhood stroke: a large-scale study of 469 childhood stroke survivors

**DOI:** 10.64898/2026.11.25.26368974

**Authors:** Joseph C. Griffis, Joel Bruss, Asmaa Mhanna, Michal Brzus, Heather J. Fullerton, Max Wintermark, Nomazulu Dlamini, Nancy K. Hills, Michael M. Dowling, Warren Lo, Timothy J. Bernard, Lori C. Jordan, Mark Mackay, Laura Lehman, Adam Kirton, Dana D. Cummings, Sarah Lee, J. Michael Taylor, Christine K. Fox, Aaron D. Boes, SCOUTS Investigators

## Abstract

The effects of stroke on the developing brain are thought to depend on age at stroke onset, but the nature of this relationship is unclear. While it has been suggested that younger age at stroke onset might facilitate recovery, there is also evidence that critical developmental periods in early childhood confer increased vulnerability to injury. Associations between age at stroke onset and childhood stroke outcomes are mixed and often inconsistent, and the roles of lesion factors such as size, laterality and location are poorly understood.

We investigated how age and lesion factors relate to chronic (6+ months post-onset) sensorimotor, expressive language, receptive language, and cognitive/behavioral impairments in 469 childhood stroke survivors enrolled in the SCOUTS study (median age at stroke=9.3 years; 57% male). We used ordinal logistic regression models to characterize associations between outcomes and age at stroke onset, left and right hemispheric lesion volumes, and years to chronic assessment, and we used voxel-based lesion-symptom correlations to evaluate associations with lesion location. We also tested whether age at stroke onset moderates relationships between lesion factors and chronic outcomes.

When considered in isolation, younger age at stroke onset was significantly associated with worse outcomes across all domains, but in multivariable models, statistically significant associations were observed only for language outcomes. Larger lesion volumes in each hemisphere were significantly associated with worse contralateral sensorimotor outcomes and with worse language and cognitive/behavioral outcomes. For language outcomes, associations with lesion volume were strongest for the left hemisphere, and older age at stroke onset amplified the detrimental effect of large left hemisphere lesions on expressive language outcomes. Lesion-symptom analyses mapped sensorimotor impairments to structures including the putamen, sensorimotor cortices, superior parietal cortex, and cortico-spinal projections. Expressive and receptive language impairments both mapped to mostly left-lateralized and partially overlapping frontal, temporal and parietal structures, with the strongest associations in the white matter underlying left posterior temporo-parietal areas. Cognitive/behavioral impairments mapped to bilateral regions associated with action mode and somato-cognitive action networks including right dorsolateral prefrontal cortex, left inferior frontal and supramarginal gyri, and inter-effector nodes of the right pre-central gyrus.

These results provide novel insights into how age at stroke onset and lesion factors contribute to childhood stroke outcomes, map domain-level impairments to specific lesion locations, and indicate that the vulnerability of left hemisphere language networks to injury may increase as language becomes more left-lateralized during development.

## Introduction

Childhood stroke, defined as strokes occurring between 29 days and 18 years of age^1^, is an uncommon but significant source of long-term morbidity and disability that can greatly reduce quality of life, lead to long-term dependence, and place substantial burdens on patients, parents, and caregivers. While some childhood stroke survivors recover without significant long-term consequences, others suffer from chronic impairments in motor, language, and/or cognitive functions^1–3^. However, the factors underlying this variability in long-term outcomes are poorly understood, posing an obstacle to accurate clinical prognostication.

The effects of stroke on the developing brain are thought to depend, in part, on the age at stroke onset^1,4,5^, but the nature of this relationship is unclear. It has been proposed that injuries occurring earlier in life may be associated with better outcomes due to enhanced plasticity and greater capacity for reorganization in the immature brain^4–8^, but there is also evidence that early-onset lesions might produce worse outcomes than lesions that occur after critical developmental periods^9,10^, and findings on this topic are mixed and often inconclusive^4,8^. For example, strokes occurring between 29 days and 6 years of age have been associated with worse impairments in cognitive flexibility, processing speed, and verbal learning compared to pediatric strokes occurring outside of this period^11^, but strokes occurring between 1 and 6 years of age have also been associated with better neuropsychological outcomes compared to strokes occurring earlier or later in childhood^12^. Interestingly, a recent study reported that strokes occurring in late childhood are associated with worse long-term outcomes than early childhood or neonatal strokes, but only after accounting for lesion factors^13^, suggesting the possibility of more complex dependencies among age at stroke onset, lesion characteristics, and long-term outcomes.

Lesion location is an important factor contributing to chronic stroke outcomes in adults^14–19^, but its association with childhood stroke outcomes is poorly understood^4,8^. Voxel-based lesion-symptom mapping (VLSM), a technique for relating outcomes after focal brain injury to the affected brain structures^20^, has been used extensively to map the anatomy underlying a wide range of post-stroke outcomes in adults^20–25^. However, lesion-symptom relationships identified in adults cannot be assumed to generalize to children^3,4^, and aside from a few recent studies that have applied VLSM to study outcomes after neonatal brain injury^26–28^, we are not aware of any studies that have applied VLSM to study chronic childhood stroke outcomes. Instead, prior pediatric lesion studies have primarily investigated how chronic outcomes relate to coarse-grained lesion features such as total infarct volume^29–31^, the affected arterial territories^32,33^ and whether the lesion affects cortical structures, subcortical structures or both, limiting the granularity of identifiable relationships. Findings regarding lesion-symptom relationships in children have been largely inconclusive, with conflicting evidence regarding basic but clinically relevant questions. For example, there are conflicting reports regarding whether lesion laterality contributes to the risk of chronic language impairments after childhood stroke^4,8,34–36^.

We therefore aimed to comprehensively characterize how age at stroke onset and lesion size, laterality and location relate to chronic domain-level neurological impairments after childhood stroke. Because the effects of brain lesions may depend on the developmental state of the brain at the time of injury^8,10^, we also aimed to evaluate whether age at stroke onset modifies the effects of lesion factors on childhood stroke outcomes. To this end, we analyzed lesion and chronic neurological outcome data obtained from a large multi-site pediatric arterial ischemic stroke cohort (*N*=469) enrolled in the Seizures and Children’s Outcome after Stroke (SCOUTS) study in what is, to our knowledge, the largest lesion-symptom mapping study of childhood stroke outcomes to date.

## Materials and methods

### Dataset

Clinical imaging and outcome data were obtained from *N*=528 children (age 29 days to 18 years) who were prospectively enrolled in the VIPS^37^ or VIPS II^38^ studies. Acute arterial ischemic stroke was confirmed by review of clinical presentation and neuroimaging data, which were independently reviewed by a board-certified pediatric vascular neurologist (H.J.F.) and neuroradiologist (M.W.), respectively. Long-term follow-up data were collected through the Seizures and Children’s Outcomes after Stroke (SCOUTS) study^39^. A flow diagram is provided in **Supplementary Figure 1**. The parent studies did not include neonatal stroke. Children with neuroimaging data sufficient for lesion segmentation were considered for inclusion in the analyses. Children were excluded if the time between stroke onset and neurological assessment was less than 6 months. A total of *N*=469 patients met inclusion criteria, with ages ranging from 29 days to 18 years. Clinical and demographic characteristics are summarized in **Table 1**.

**Table 1.** Clinical and demographic characteristics (N=469)

| **Demographic Characteristics** |  |
|---|---|
| Stroke Onset Age in Years, median (IQR) | 9.3 (11.02) |
| Years to Chronic Assessment, median (IQR) | 3.32 (4.26) |
| Male, *n* (%) | 266 (57) |
| Race, *n* (%) |  |
| White | 320 (68) |
| Black | 58 (12) |
| Asian | 22 (5) |
| Other | 69 (15) |
| **Stroke Classifications, *n* (%)** |  |
| Idiopathic | 25 (5) |
| Cardioembolic | 18 (4) |
| Definite Arteriopathy | 177 (38) |
| Possible Arteriopathy | 92 (20) |
| No Arteriopathy | 157 (33) |
| **Comorbidities, *n* (%)** |  |
| Chronic Heart Disease | 69 (15) |
| Endocarditis | 5 (1) |
| Head Trauma | 45 (10) |
| Meningitis | 21 (5) |
| Sickle Cell | 16 (3) |
| Systemic Sepsis | 18 (4) |
| **Lesion Laterality, *n* (%)** |  |
| Left | 180 (38) |
| Right | 193 (41) |
| Bilateral | 96 (21) |
| **Lesion Volume (mean ± SD)** |  |
| Total | 63999±107245 |
| Left Hemisphere | 52939±85387 |
| Right Hemisphere | 53304±100620 |

Note: Lesion volume estimates are computed as counts using 1mm isotropic voxels. Means and standard deviations for left and right hemispheric lesion volumes were computed only for patients with damage to each hemisphere.

### Outcome measures

Outcomes were measured using the Pediatric Stroke Outcome Measure (PSOM)^40^, a validated measure of functional outcome based on clinical neurologic assessment that scores deficits in five domains: right and left sensorimotor, expressive language, receptive language, and cognitive/behavior. If PSOM data were not available, outcomes were measured using the Recurrence and Recovery Questionnaire (RRQ)^41^, a parent-administered assessment that correlates strongly with the PSOM^41^. PSOM and RRQ are scored on a 4-point ordinal scale. Higher scores indicate greater impairment.

### Standard protocol approvals, registrations, and patient consents

The SCOUTS study was approved by an Institutional Review Board (IRB) at Vanderbilt University Medical Center, which served as the IRB of record for all participating domestic sites. Local IRB approved the study at all participating sites outside the United States in the SCOUTS study and at each enrolling site for the VIPS and VIPSII studies. This analysis was approved by the University of Iowa Institutional Review Board (Project ID 201705278). Written informed consent was obtained from all participants or parents of children, as applicable, in accordance with the Declaration of Helsinki. In some instances, chart review was performed for follow-up data in participants that had previously consented to research, and the need for re-consenting participants was waived as the research was deemed minimal risk. Follow-up data were collected through chart review with a waiver of consent for minimal risk research, or through standardized parental questionnaires.

### Neuroimaging data

Neuroimaging data collection and image processing steps are described in full detail elsewhere^39^, and are only briefly described here. Typical pediatric stroke MRI protocols included T1, FLAIR, gradient echo, diffusion weighted imaging (DWI), and apparent diffusion coefficient (ADC) sequences. Raw imaging DICOMs were sent from the participating sites to the IPSS Neuroimaging Core at the Hospital for Sick Children in Toronto^42^, and from there to the University of Iowa IPSS Imaging Core. Lesions were manually traced in native space using FSL, with primary reliance on DWI and ADC. The native MRI and co-registered lesion mask were aligned to an MNI152 template brain via linear and nonlinear registrations implemented in ANTs. SynthSR was used to upsample and normalize clinical images to 1mm isotropic resolution prior to registration^43^. Enantiomorphic normalization was used to improve registration for unilateral lesions^44^, and cost-function masking^45^ was used for bilateral lesions. All lesion segmentations and registrations were reviewed by experts, including A.D.B. and J.B., and any errors were manually corrected. Investigators involved in lesion tracing and editing were blind to behavioral outcomes. Lesion volumes were estimated as 1mm isotropic voxel counts in template space to remove variability due to factors such as head size or brain volume. Lesion segmentations were resampled to 2mm isotropic voxel resolution prior to VLSM analyses.

### Statistical analyses

#### Ordinal logistic regression models of chronic neurological outcomes

To characterize bivariate relationships between each outcome and age at stroke onset, we first fit single-variable ordinal logistic regression models that included only age at stroke onset as a predictor. We then fit multivariable ordinal logistic regression models that included age, lesion size, and lesion laterality as predictors of interest. Time from stroke onset to chronic neurological assessment was also included as a covariate in these models and in subsequent VLSM analyses since it has the potential to influence associations among the variables of interest^8^. Time since stroke was used rather than age at assessment because the latter was colinear with age at stroke onset (*r*=0.83, *P*<0.001).

To test whether the effects of extensive damage to each hemisphere on chronic outcomes might depend on age at stroke onset, we defined interaction terms between each of the hemispheric lesion volume variables and age at stroke onset, and we used a bootstrapped (1000 iterations per outcome) Bayesian Information Criterion (BIC) model selection procedure to identify the most likely model given the data^46,47^. Specifically, we fit four nested models for each outcome that included the same set of predictors described above, but differed with respect to the included interaction terms: Model 1 included no interaction terms and corresponded to the “base” model, Model 2 included only an interaction term for right hemispheric lesion volume, Model 3 included only an interaction term for left hemispheric lesion volume, and Model 4 included separate interaction terms for left hemispheric and right hemispheric lesion volumes (i.e. “full” model). The model that was selected most frequently across bootstrap iterations was selected as the “final” model for interpretation, although for completeness, we also explored the moderation effects from the “full” model (i.e. Model 4) for each outcome.

We interpreted individual predictor effects in each multivariable model if the omnibus likelihood ratio test was statistically significant after Bonferroni-Holm family-wise error (FWE) correction across omnibus tests for all outcomes at P_FWE_<0.05. For completeness, we report both per-comparison and Bonferroni-Holm P-values for each predictor in each model, with Bonferroni-Holm correction applied across all predictors included within each model. We also report 95% CIs on the adjusted cumulative odds ratios (aORs) for each standardized predictor. Selected models that included interaction effects were followed up with conditional effects analyses that estimated aORs and 95% CIs for the effect of a 1 standard deviation increase in the lesion volume of the relevant hemisphere at standardized age levels defined at -1.5, -1, 0, and 1 standard deviations from the average age, corresponding to ∼3.5m, ∼3y, ∼9y, and ∼15y of age.

### Voxel-based lesion-symptom mapping

We conducted mass-univariate voxel-based lesion-symptom mapping (VLSM) analyses using the Iowa Brain Behavior Modeling Toolkit^48^, available at (https://github.com/jcgriffis/ibb_toolkit), running in MATLAB R2025b (The MathWorks), computing non-parametric Spearman correlations at each voxel to test for associations between voxel lesion status and each outcome. Mass-univariate VLSM allowed for the use of non-parametric statistics (i.e. rank correlations) that are appropriate for evaluating monotonic associations with ordinal outcomes that may be non-linear. We note that large-scale comparisons of mass-univariate and multivariate VLSM approaches have not identified a consistent advantage for multivariate approaches in inferential analyses^49^, and both approaches are similarly susceptible to result displacement when not accounting for lesion size^50,51^.

Voxels were analyzed if they were lesioned in at least 5 patients^51^. Analyses were conducted using both full and partial Spearman correlations to illustrate the raw lesion-symptom relationships and to provide clarity regarding the effects of covariate control on the results^52^. Partial Spearman correlation analyses included age at stroke onset, time from stroke onset to neurological evaluation, and left and right hemispheric lesion volumes as covariates since they have the potential to influence lesion-symptom associations^8^. We chose the conservative approach of separately modeling left and right hemispheric lesion volumes to allow for the possibility that associations between lesion volume and chronic outcomes might be lateralized, in which case covarying total lesion volume would misestimate the true effect of lesion volume in each hemisphere.

Statistical significance was determined using permutation testing (5000 permutation iterations). Permutation testing for partial correlation analyses used the Freedman-Lane procedure^53^. Whole-brain statistical thresholds were determined using the continuous family-wise error control (cFWE) procedure developed and validated for mass-univariate VLSM^54^. This procedure identifies FWE-corrected thresholds that control the probability of at least *v* false positive voxels across the whole brain at the nominal alpha level (i.e. 0.05), where *v* is a user-selected suprathreshold voxel count parameter. Here, we used *v*=100. Following the procedures described by Mirman and colleagues^54^, we also calculated the post-hoc “effective false discovery rate” (eFDR) at the selected *v=*100 threshold, reflecting the proportion of surviving voxels that are expected to be false positives given the selected *v* threshold, and the number of suprathreshold voxels. For example, if 2000 voxels survived at a *v*=100 threshold, the eFDR would be 5% (i.e. eFDR=100/2000). Results were considered statistically significant if the post-hoc eFDR was less than or equal to 5%.

For the primary LSM of sensorimotor outcomes reported in the main text, all lesions were flipped across the midline to render them mirror-symmetric, and the sensorimotor outcome variable was defined as the sensorimotor domain score corresponding to the body side contralateral to the stroke lesion; for patients with bilateral lesions, the sensorimotor domain with the highest impairment score was used, and flipped lesions included all lesion sites in both hemispheres. Separate analyses were also performed for each outcome using the original un-flipped lesions.

### Voxel-based moderation analyses of stroke onset age and lesion status

To interrogate whether voxel-level lesion-symptom associations varied depending on age at stroke onset, we performed voxel-wise moderation analyses by fitting ordinal logistic regression (proportional odds) models at each voxel that included (1) an interaction term between age at stroke onset and that voxel’s binary lesion status, and (2) the main effects of each variable, along with left and right hemispheric lesion volumes and the time interval from stroke onset to chronic assessment as covariates.

Permutation testing (1000 iterations) was used to determine the statistical significance of the moderation effects using a permutation testing approach for logistic regression effects in the presence of covariates^55^. We used a smaller number of permutations than for the LSM analyses due to the substantially higher computational time required for permutation analyses of voxel-wise ordinal logistic regressions, but prior work indicates that as few as 500 permutations is sufficient to produce stable and reliable *P*-values when computational demands are a concern^56^. The cFWE procedure was used to control the whole-brain FWE for each analysis at 0.05 with *v=*100. Results were considered statistically significant if the post-hoc eFDR was less than or equal to 5%. Statistically significant results were characterized via conditional effects analyses for peak voxels, analogous to those described for the multivariable ordinal logistic regression models above, evaluating the effects of a binary change in lesion status (i.e. 0 to 1) at each level of age.

## Results

### Lesion, outcome, onset age, and assessment interval distributions

Lesion coverage spanned the majority of the cerebrum, with the exception of some medial structures primarily in the left hemisphere (**Figure 1A**). Lesions most frequently affected the bilateral putamen and adjacent structures, and to a lesser degree, lateral frontal, parietal, and temporal lobe structures, with slightly reduced frequencies and coverage in the left relative to right hemisphere.

**Figure 1.**
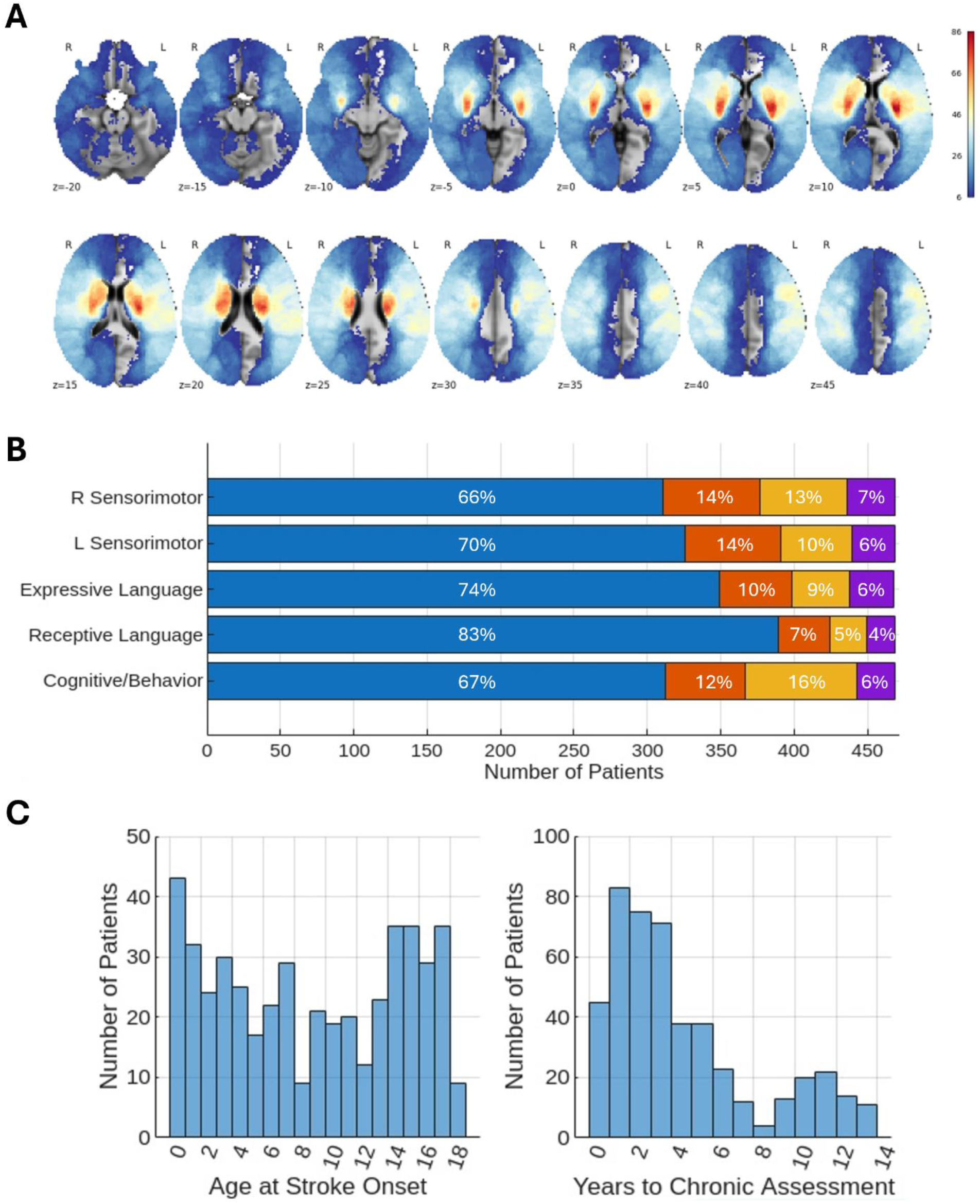
**A.** Lesion frequency maps. Colors reflect the number of patients with lesions affecting each voxel. Voxels affected in at least 5 patients are shown. **B.** PSOM/RRQ score distributions for each domain (N=469). The number of patients (x-axis) with each PSOM/RRQ score (blue = 0, orange = 0.5, yellow = 1, purple = 2) is shown for each PSOM/RRQ domain (y-axis). **C.** Distribution of ages at stroke onset (left) and distribution of years from onset to chronic neurological assessment (right).

Across outcomes, some level of impairment (i.e. PSOM/RRQ > 0) was observed in ∼17-34% of patients, with impairments being most common for sensorimotor and cognitive/behavioral outcomes, and least common for the receptive language outcome (**Figure 1B**). The majority of chronic impairments tended to be mild or moderate, with fewer than 10% of patients exhibiting severe impairments in any outcome domain. Stroke onset ages effectively spanned the full range of early and late childhood stroke (**Figure 1C**). Chronic assessments were most frequently obtained within the first 4 years of stroke onset, and 98% (*n*=459) of included patients were 2 years or older at the time of assessment.

### Effects of lesion size, laterality, and age at stroke onset

#### Associations between age at stroke onset and chronic impairments

Children with more severe impairment in a given outcome domain tended to have earlier stroke onsets than those with less severe impairments (**Supplementary Figure 2A**), and younger age at stroke onset was consistently associated with worse chronic outcomes across all outcome domains in the single-variable models, with the strongest associations observed for the receptive and expressive language outcomes (**Supplementary Figure 2A**). Correlations among predictor variables are shown in **Supplementary Figure 2B**.

#### Multivariable models of chronic impairments

The selected multivariable model for each outcome, determined according to the bootstrapped BIC model comparison procedure, is shown in **Figure 2A**. Omnibus likelihood ratio tests were statistically significant for all selected multivariable models (all χ^2^>=33.92, *P_FWE_* < 0.001), supporting interpretation of the individual predictor effects (**Table 2**). Model 1 was selected for all outcomes except for expressive language, where Model 3 was selected.

**Figure 2.**
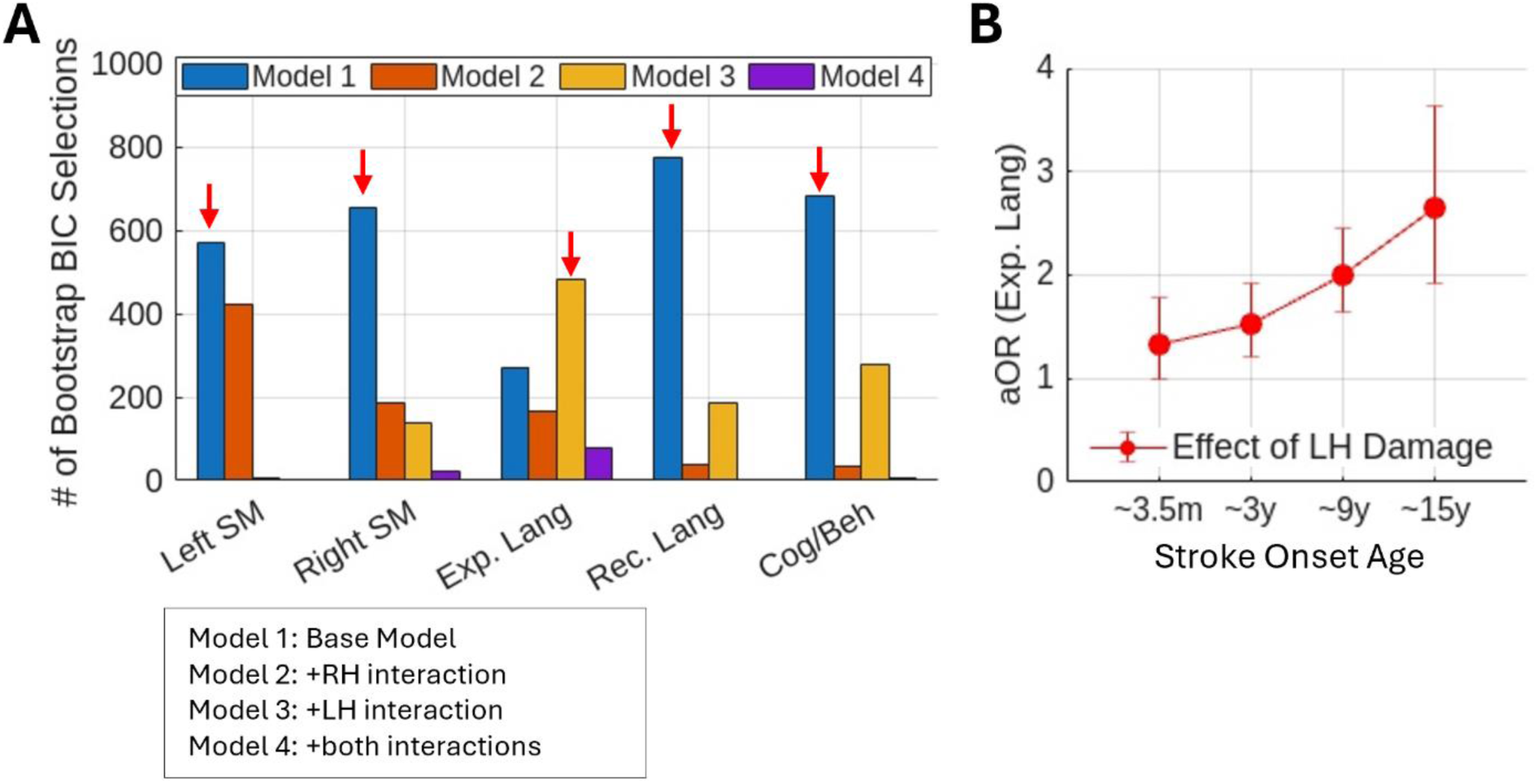
Model selection and moderation analysis results. **A.** For each outcome, the bars show the number of times each candidate model was selected during the bootstrapped BIC model selection procedure (1000 iterations). The “final” model for each outcome, corresponding to the model selected most frequently during the bootstrapped BIC model selection procedure, is identified by a red arrow. **B.** Conditional effects of a 1SD increase in left hemispheric lesion volume on expressive language outcomes are shown with 95% CIs at -1.5, -1, 0, and 1 standard deviations from the mean stroke onset age (∼3.5m, ∼3y, ∼9y, and ∼15y, respectively).

**Table 2.** Multivariable Ordinal Logistic Regression Models.

| **Left SM** | **Std. Beta** | **aOR** | **95% CI** | **t-statistic** | ***P*-value** | ***P<sub>FWE</sub>*** |
|---|---|---|---|---|---|---|
| Age at Stroke | -0.09 | 0.91 | 0.74, 1.13 | -0.82 | 0.41 | 0.41 |
| Years to Test | 0.15 | 1.17 | 0.96, 1.42 | 1.54 | 0.12 | 0.24 |
| Lesion Volume (LH) | -0.48 | 0.62 | 0.44, 0.87 | -2.74 | 0.006 | 0.02 |
| Lesion Volume (RH) | 0.75 | 2.12 | 1.70, 2.65 | 6.70 | 2.13e-11 | <0.001 |
| **Right SM** | **Std. Beta** | **aOR** | **95% CI** | **t-statistic** | ***P*-value** | ***P<sub>FWE</sub>*** |
| Age at Stroke | -0.20 | 0.82 | 0.67, 1.01 | -1.88 | 0.06 | 0.18 |
| Years to Test | 0.02 | 1.02 | 0.84, 1.24 | 0.19 | 0.85 | 0.85 |
| Lesion Volume (LH) | 0.84 | 2.31 | 1.89, 2.83 | 8.13 | 4.43e-16 | <0.001 |
| Lesion Volume (RH) | -0.15 | 0.86 | 0.70, 1.07 | -1.35 | 0.18 | 0.36 |
| **Expressive Lang** | **Std. Beta** | **aOR** | **95% CI** | **t-statistic** | ***P*-value** | ***P<sub>FWE</sub>*** |
| Age at Stroke | -0.29 | 0.75 | 0.60, 0.95 | -2.41 | 0.02 | 0.04 |
| Years to Test | 0.16 | 1.17 | 0.95, 1.44 | 1.49 | 0.14 | 0.14 |
| Lesion Volume (LH) | 0.70 | 2.01 | 1.65, 2.45 | 6.87 | 6.25e-12 | <0.001 |
| Lesion Volume (RH) | 0.34 | 1.41 | 1.17, 1.69 | 3.64 | 0.0003 | 0.001 |
| LH Volume x Age | 0.28 | 1.32 | 1.09, 1.60 | 2.78 | 0.005 | 0.02 |
| **Receptive Lang** | **Std. Beta** | **aOR** | **95% CI** | **t-statistic** | ***P*-value** | ***P<sub>FWE</sub>*** |
| Age at Stroke | -0.49 | 0.61 | 0.47, 0.80 | -3.53 | 0.0004 | 0.001 |
| Years to Test | 0.07 | 1.07 | 0.84, 1.37 | 0.57 | 0.57 | 0.57 |
| Lesion Volume (LH) | 0.51 | 1.67 | 1.38, 2.02 | 5.31 | 1.13e-07 | <0.001 |
| Lesion Volume (RH) | 0.24 | 1.27 | 1.05, 1.54 | 2.45 | 0.014 | 0.03 |
| **Cog/Behavioral** | **Std. Beta** | **aOR** | **95% CI** | **t-statistic** | ***P*-value** | ***P<sub>FWE</sub>*** |
| Age at Stroke | -0.14 | 0.86 | 0.71, 1.05 | -1.45 | 0.15 | 0.15 |
| Years to Test | 0.21 | 1.23 | 1.02, 1.48 | 2.17 | 0.03 | 0.06 |
| Lesion Volume (LH) | 0.33 | 1.39 | 1.17, 1.65 | 3.71 | 0.0002 | <0.001 |
| Lesion Volume (RH) | 0.30 | 1.35 | 1.13, 1.61 | 3.34 | 0.0008 | 0.002 |

**Note:** Beta coefficients and adjusted cumulative odds ratios (aOR) for continuous predictors are based on standardized variables and represent effects associated with a 1 standard deviation increase in the predictor variables. aOR values > 1 indicate higher odds of impairment, while aOR values < 1 indicate lower odds of impairment. 95% CIs are provided for aORs, along with *t*-statistics and *P*-values.

In the multivariable models, younger age at stroke onset remained consistently associated with worse outcomes across domains, but statistically significant associations were only observed for the language outcomes. Larger lesion volumes in each hemisphere were significantly associated with worse sensorimotor outcomes for the contralateral body side, and with worse language and cognitive behavioral outcomes. For the receptive language model, the standardized effect was greater for left relative to right hemispheric lesion volume. For the expressive language model, the effect of left hemispheric lesion volume was moderated by age at stroke onset. Follow-up conditional effects analyses indicated that the detrimental effect of extensive left hemispheric damage on expressive language outcomes increased with older age at stroke onset (**Figure 2B**). For completeness, conditional effects plots for the moderation effects from the “full” models (i.e. Model 4) of each outcome are shown in **Supplementary Figure 3**.

### Lesion-symptom mapping of chronic impairments

#### Contralateral sensorimotor outcome

The strongest and most robust associations were observed in the putamen, internal capsule, pre-central gyrus, post-central gyrus, inferior parietal lobe, superior parietal cortex, insula, superior temporal gyrus, and portions of the deep white matter containing both projection and association pathways (**Figure 3**; **Supplementary Figure 4**). Similar, but sparser, results were obtained when separately modeling left vs. right sensorimotor domains using the original (i.e. unflipped) lesion masks (**Supplementary Figure 5**), consistent with the expectation that rendering the lesions mirror symmetric would increase statistical power. The partial correlation analysis of the right sensorimotor domain also identified significant associations with lesions affecting the right cerebellar vermis and lobule VI (**Supplementary Figure 5**).

**Figure 3.**
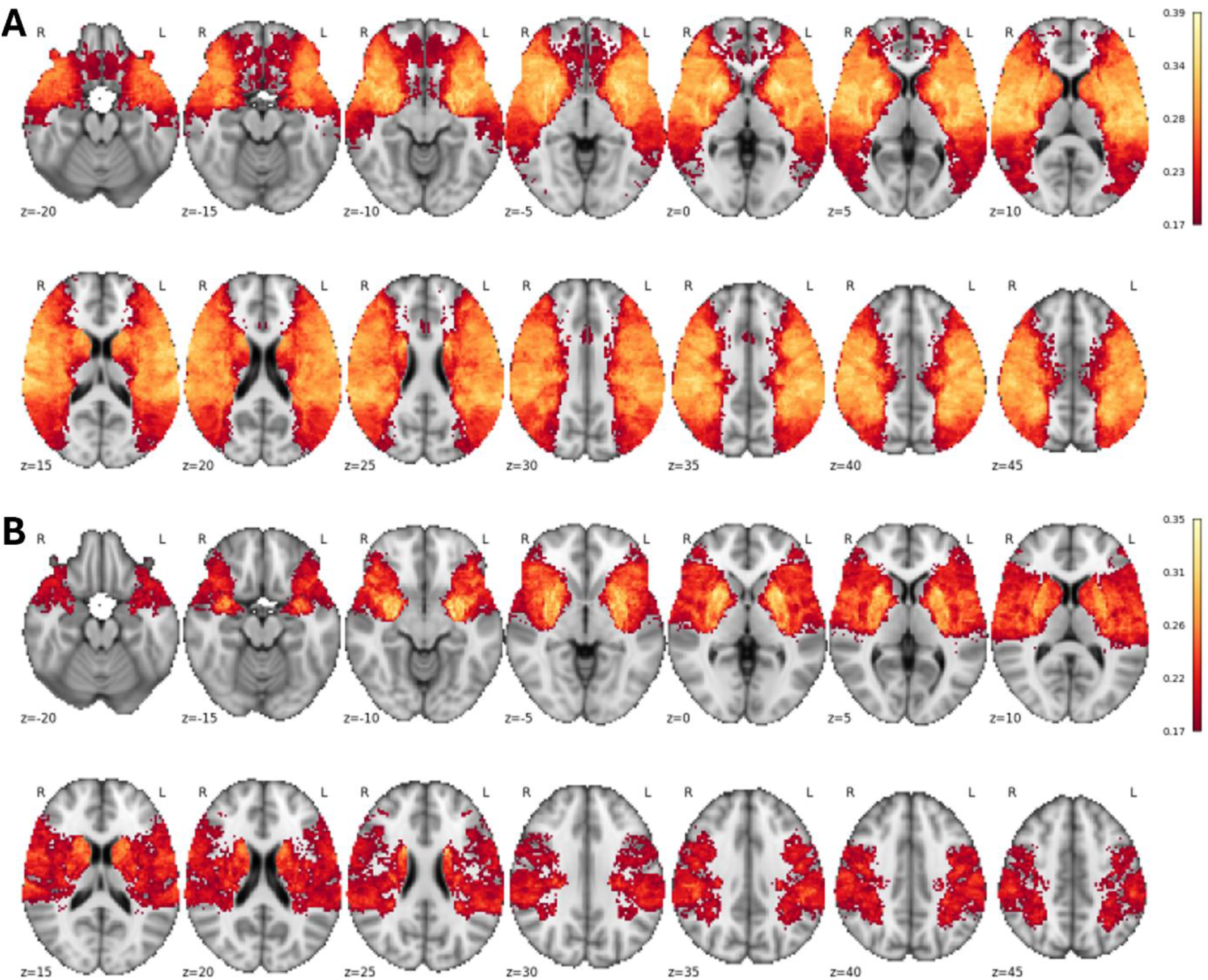
Lesion-symptom mapping results for the contralateral sensorimotor outcome. **A.** Full correlation results. Colors reflect full Spearman rank correlations between lesion status and the contralateral sensorimotor outcome. Results are statistically significant at *_PFWE_*< 0.05 for *v*=100 voxels, with a post-hoc effective FDR of 0.001. **B.** Partial correlation results. Colors reflect partial Spearman rank correlations between lesion status and the contralateral sensorimotor outcome, covarying age at stroke onset, time from stroke onset to neurological assessment, and left and right hemispheric lesion volumes. Results are statistically significant at *P_FWE_*< 0.05 for *v*=100 voxels, with a post-hoc effective FDR of 0.002. Note – the map is mirror-symmetric due to the use of symmetrically flipped lesion masks. Separate maps for the left and right sensorimotor outcomes are provided in **Supplementary Figure 5**.

#### Expressive language outcome

The strongest and most robust associations were observed in the left posterior superior/middle temporal cortices, the supramarginal gyrus, the anterior superior temporal gyrus, and portions of the deep white matter containing primarily left-hemispheric association and projection pathways (**Figure 4; Supplementary Figure 4**). Statistically significant associations were also observed in a set of primarily left-hemispheric structures, including the inferior frontal cortex, as well as in some right hemispheric structures including the posterior superior temporal gyrus, lateral posterior temporooccipital cortex, precentral gyrus, and dorsolateral prefrontal cortex.

**Figure 4.**
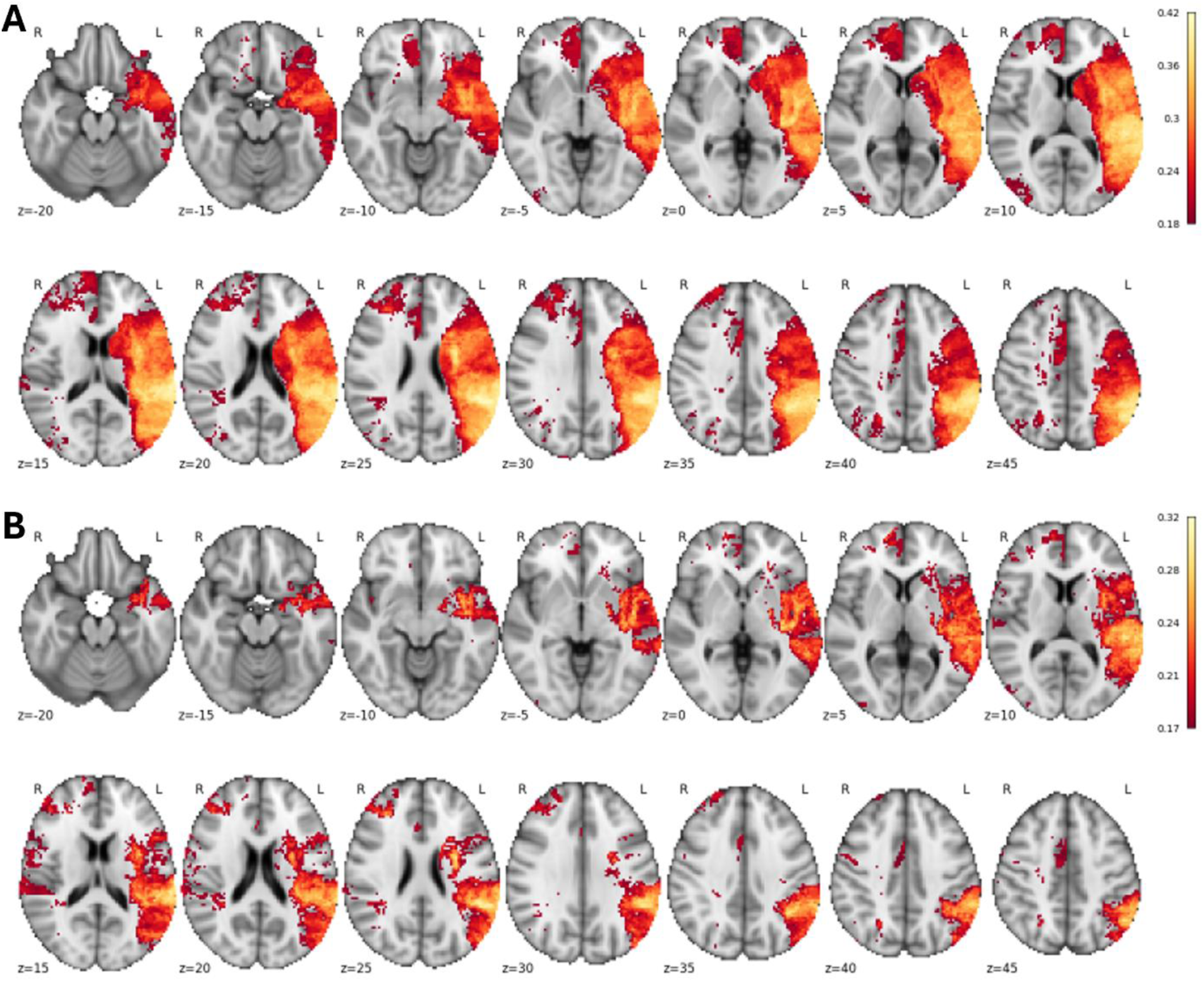
Lesion-symptom mapping results for the expressive language outcome. **A.** Full correlation results. Colors reflect full Spearman rank correlations between lesion status and the expressive language outcome. Results are statistically significant at *P_FWE_* < 0.05 for *v*=100 voxels, with a post-hoc effective FDR of 0.002. **B.** Partial correlation results. Colors reflect partial Spearman rank correlations between lesion status and the expressive language outcome, covarying age at stroke onset, time from stroke onset to neurological assessment, and left and right hemispheric lesion volumes. Results are statistically significant at *P_FWE_*< 0.05 for *v*=100 voxels, with a post-hoc effective FDR of 0.005.

#### Receptive language outcome

The strongest and most robust associations were observed in the deep white matter underlying the posterior superior temporal gyrus and supramarginal gyrus, consistent with the expected trajectories of the arcuate and, to a lesser extent, inferior fronto-occipital fasciculi (**Figure 5**; **Supplementary Figure 4**). Statistically significant associations were also observed with primarily left-lateralized regions in the frontal, temporal, and parietal lobes, including inferior frontal cortex, posterior middle and inferior temporal cortices, and the anterior temporal lobe, and in the right prefrontal white matter, right dorsolateral prefrontal cortex, and right inferior temporo-occipital cortex.

**Figure 5.**
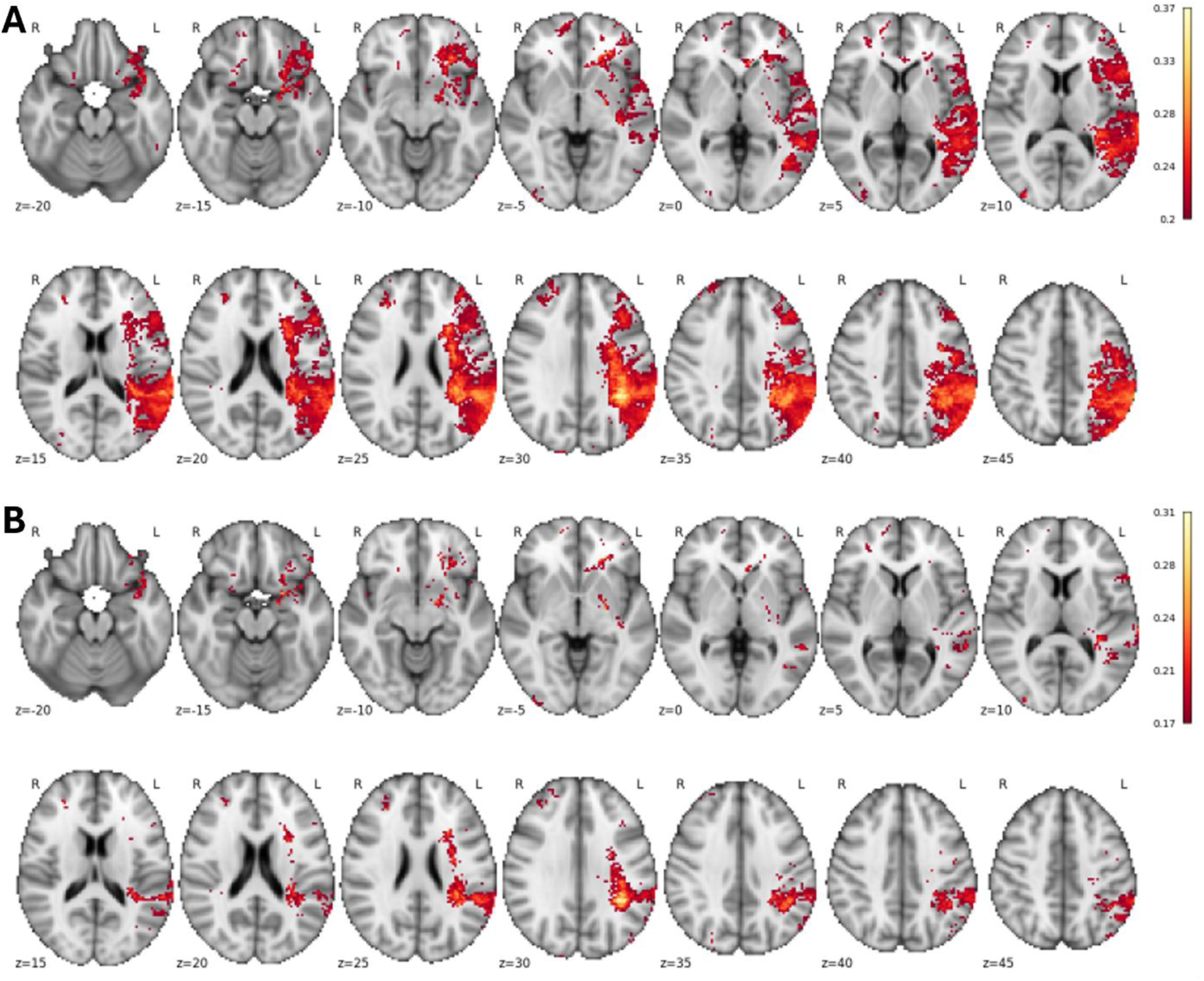
Lesion-symptom mapping results for the receptive language domain. **A.** Full correlation results. Colors reflect full Spearman rank correlations between lesion status and the receptive language outcome. Results are statistically significant at *P_FWE_*< 0.05 for *v*=100 voxels, with a post-hoc effective FDR of 0.005. **B.** Partial correlation results. Colors reflect partial Spearman rank correlations between lesion status and the receptive language outcome, covarying age at stroke onset, time from stroke onset to neurological assessment, and left and right hemispheric lesion volumes. Results are statistically significant at *P_FWE_*< 0.05 for *v*=100 voxels, with a post-hoc effective FDR of 0.02.

#### Cognitive and behavioral outcome

Statistically significant results were only obtained using the full correlation approach, and observed associations were comparatively sparse, with the strongest associations observed in the right dorsolateral prefrontal cortex and right pre-central gyrus, and significant clusters in portions of the basal ganglia, left inferior frontal cortex, left supramarginal gyrus, and bilateral insulae (**Figure 6**). Notably, many of the cortical clusters appeared to correspond to portions of the action mode network (AMN)^57^, and the peaks of the pre-central gyrus cluster showed close correspondence to the inter-effector nodes of the somato-cognitive action network (SCAN)^58^, which is closely related to the AMN (**Supplementary Figure 6**). The peak in the right dorsolateral prefrontal cortex also showed close correspondence to a lesion site previously linked to dysexecutive personality disturbances in adults^59^.

**Figure 6.**
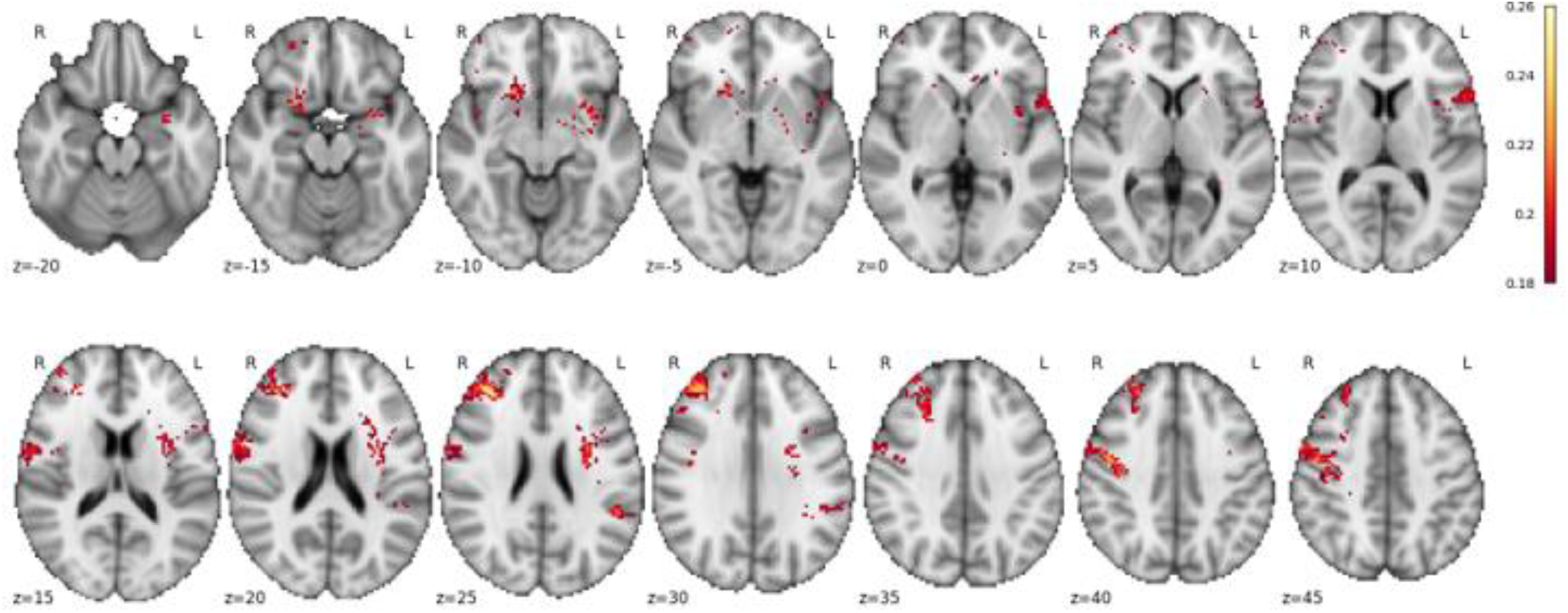
Lesion-symptom mapping results for the cognitive and behavioral outcome. Full correlation results. Colors reflect full Spearman rank correlations between lesion status and the cognitive and behavioral outcome. Results are statistically significant at *P_FWE_*< 0.05 for *v*=100 voxels, with a post-hoc effective FDR of 0.03. No statistically significant results were obtained when covarying age at stroke onset, time from stroke onset to neurological assessment, and left and right hemispheric lesion volumes.

#### Moderating effects of age on voxel-level lesion-symptom associations

We did not identify any statistically significant voxel-level moderation effects for any outcome domain (all post-hoc FDRs > 0.35), even when using a relaxed *v* threshold (*v=*1000, all post-hoc FDRs > 0.33) or when applying more liberal voxel-wise FDR correction^60^ to parametric *P*-values obtained from the voxel-level ordinal logistic regression models. Unthresholded statistical maps obtained from these analyses are provided in **Supplementary Figure 7.**

## Discussion

Here, we aimed to characterize how age at stroke onset, lesion size and laterality, and lesion location relate to chronic neurological outcomes after childhood stroke. Younger age at stroke onset showed consistent associations with worse outcomes across domains when modeled in isolation. When age was included in the multivariable models, we observed domain-specific associations with worse language outcomes. Larger lesions in each hemisphere were associated with worse sensorimotor outcomes for the contralateral body side, and with worse cognitive/behavioral outcomes. While extensive damage to both the left and right hemispheres was associated with worse language outcomes, associations were strongest for the left hemisphere, and the risk of poor expressive language outcomes conferred by extensive left hemispheric damage increased with older age at stroke onset. Beyond characterizing these associations, we also generated the most comprehensive set of pediatric lesion-symptom maps from the largest pediatric lesion sample to date, made possible by a large international consortium collecting data for over a decade, which we will make available as a resource for clinicians and researchers. Together, these results provide novel insights into how chronic impairments after childhood stroke relate to age at stroke onset and to the underlying lesion anatomy.

### Associations between age at stroke onset and chronic outcomes

There are opposing views on how age at stroke onset relates to outcomes after pediatric stroke. Strokes occurring earlier in life might allow for greater plasticity and more effective compensation that could facilitate recovery^5,6,8^, but strokes occurring in early childhood have been associated with worse outcomes relative to neonatal strokes^11,33^, suggesting a period of increased vulnerability in early childhood^8,10^. In our sample, younger age at stroke onset was generally associated with worse outcomes across domains (**Supplementary Figure 2A**), but when age was modeled jointly with lesion factors, it only showed significant associations with language outcomes (**Table 2**). This suggests that age at onset, lesion characteristics, and chronic outcomes are interdependent. Indeed, age at onset was weakly but significantly correlated with both left and right hemispheric lesion volumes in our sample (**Supplementary Figure 2B**), suggesting that the observed associations between age at onset and chronic outcomes were partially driven by correlated lesion laterality and volume effects. A recent study reported the opposite pattern, i.e. that older age at stroke onset increased the risk of poor outcomes, but only when onset age was modeled jointly with lesion characteristics^13^, although several differences complicate direct comparisons; for example, that study featured a substantially smaller patient sample than the current study (i.e. *N*=285) and examined stroke onset age using relatively large age bins (i.e. neonatal, 1 month to 5 years, and 5+ years). Even so, both studies point more generally towards the existence of dependencies among age at stroke onset, lesion characteristics, and chronic outcomes, such that the age-outcome relationships observed in a given sample might depend on how lesion characteristics are distributed across stroke onset ages, highlighting the importance of modeling lesion factors when evaluating age-outcome relationships.

Notably, we found that the detrimental effect of larger left hemisphere lesions on expressive language outcomes increased with older age at stroke onset (**Figure 2B**). Similar trends were observed for receptive language, and right hemisphere lesions also showed a similar trend in the opposite direction both when exploring the moderation effects from the “full” moderation models (**Supplementary Figure 3**) and when inspecting the unthresholded maps from the voxel-wise moderation analyses (**Supplementary Figure 7)**. Together, these observations suggest the hypothesis that the detrimental effects of extensive left vs. right hemisphere damage on expressive language outcomes may shift in opposite directions as language systems become increasingly left-lateralized during development^61,62^. However, despite our large sample size, the only statistically significant moderation effect was that of age at stroke onset on the relationship between left hemispheric lesion volume and expressive language outcomes, and while the exploration of subthreshold and statistically non-significant effects is useful for hypothesis generation, these hypotheses require confirmation in larger patient samples.

### Lesion-symptom mapping of sensorimotor outcomes

Impairments on the sensorimotor PSOMs have been associated with poor performance on neuropsychological tests measuring visuomotor coordination and fine motor skills^40^. We observed robust anatomical associations with sensorimotor outcomes that localized primarily to regions such as the putamen, internal capsule, primary somatosensory/motor areas, and white matter locations consistent with the trajectories of cortico-striatal, cortico-spinal, and other projection pathways (**Figure 3, Supplementary Figure 4A**). These results appear broadly consistent with prior work identifying lesion correlates of motor and somatosensory deficits^63–67^ and functional outcomes^68^ in adult stroke patients and in survivors of neonatal stroke^27,28,69,70^, suggesting that lesion locations associated with chronic sensorimotor impairments may be relatively consistent across the lifespan^4^.

### Lesion-symptom mapping of language outcomes

Impairments on the expressive language PSOM have been associated with poor performance on expressive vocabulary and category fluency tests, while impairments on the receptive language PSOM have been associated with poor performance on receptive vocabulary tests^40^. While factors such as lesion laterality have not been consistently associated with language outcomes after pediatric stroke^4,8,34^, we found that chronic language impairments were more strongly associated with the extent of damage to the left hemisphere (**Table 2**, **Supplementary Figure 3**) and showed robust voxel-level associations with lesions affecting predominantly left-lateralized structures (**Figures 4-5**).

Classical models of language organization posited separate cortical modules for speech comprehension and production in the posterior temporal and inferior frontal cortices, respectively, but converging lines of evidence indicate that these processes depend on dynamic interactions across distributed fronto-temporo-parietal networks that are organized via dorsal and ventral processing streams^19,71–74^. Our results are generally consistent with this view^75^, finding no clear anterior/posterior distinction between lesion sites associated with expressive and receptive language deficits, and impairments in both sub-domains localized to a set of primarily left-lateralized regions including the posterior superior and middle temporal cortices, anterior temporal lobe, supramarginal gyrus, inferior frontal cortex, and deep white matter locations consistent with the trajectories of white matter pathways subserving the dorsal and ventral processing streams such as the arcuate, superior longitudinal, and inferior fronto-occipital fasciculi^72,76,77^ (**Supplementary Figure 4**).

Notably, both expressive and receptive language outcomes showed the strongest associations with lesions affecting posterior temporo-parietal structures and the underlying deep white matter (**Figures 4**–**5**). There is evidence that these regions correspond to language network hubs^78,79^, and white matter pathways supporting both the dorsal and ventral processing streams converge in the underlying deep white matter^80–82^. Accordingly, lesions affecting these structures have been associated with broad and enduring language impairments in adults^19,80,82–84^, including language comprehension deficits^85^ and poor recovery of impaired naming abilities^86^, and with abnormal fMRI activation patterns during language tasks^79,87^, suggesting that damage may disrupt distributed processing throughout the broader language network^79^.

Developmental factors could also explain why damage to these regions is associated with poor language outcomes in children. Language network development involves two general stages, with the first stage corresponding to the development of bottom-up systems starting in the posterior temporal cortices and extending to the anterior temporal lobe across the first 3 years of life, and the second stage culminating in the development of capabilities for sentence-level processing of complex syntax via fronto-temporal pathways that are solidified during adolescence^88^. Lesions affecting posterior temporal structures and their connections during the first stage might impede the establishment of foundational bottom-up pathways in the temporal lobe, while lesions affecting the same structures during the second stage might disrupt the establishment and refinement of top-down fronto-temporal pathways, suggesting that critical periods of vulnerability may exist in both early and late childhood. Consistent with this view, a recent study of 53 children with neonatal stroke found that damage to posterior temporo-parietal regions was associated with increased need for speech therapy at 2 years of age^28^, suggesting that even very early damage to these structures can substantially impede normal language development.

### Lesion-symptom mapping of cognitive and behavioral outcomes

Impairments on the cognitive/behavioral PSOM have been associated with lower full-scale IQ and with impairments in self-control, metacognition, and, most prominently, behavioral regulation^40^. Unlike the language outcomes, no clear laterality effect was observed for cognitive/behavioral outcomes, with extensive damage to each hemisphere independently conferring similar risk of poor outcomes (**Table 2**). While statistically significant results were only obtained for the full correlation analyses (**Figure 6**), the spatial topographies of the unthresholded maps obtained from the full vs. partial correlation analyses were highly similar (spatial *r*=0.93), indicating that the covariates had little impact on the spatial pattern of lesion-symptom associations but primarily served to attenuate the observed correlations.

Impairments localized, in part, to bilateral frontal, insular, and inferior parietal areas associated with networks involved in cognitive control processes such as the cingulo-opercular/AMN and frontoparietal control networks^89,90^ (**Figure 6**). Damage to some of the implicated structures, such as the inferior frontal cortex, supramarginal gyrus, insula, and precentral/postcentral gyri has also been previously associated with impairments on subtests of the WAIS in adults^23^, consistent with prior studies linking the cognitive and behavioral PSOM to measures of full-scale IQ^40^. Local peaks within the right pre-central gyrus cluster also showed close correspondence to inter-effector nodes of the SCAN (**Supplementary Figure 6**), which is thought to be involved in integrating motor/autonomic control with action planning, motivation, and volition^58^. These regions are also functionally connected with the AMN^58^, consistent with involvement in the regulation, modulation, and coordination of motor outputs to support goal-oriented behavior, and the right dorsolateral prefrontal cluster both overlapped with the AMN and was adjacent to a lesion site that has previously been associated with dysexecutive personality disturbances in adults^59^.

### Limitations

Our study has limitations that should be considered when interpreting the results and planning future studies in this area.

First, while the PSOM/RRQ are commonly used and validated tools for measuring outcomes after pediatric stroke, they are also very coarse-grained both in terms of the actual measurement scale (i.e. 4-point ordinal scale) and in terms of the granularity of the domain-level assessments. For example, the sensorimotor outcome scores do not differentiate among distinct types of sensorimotor deficits and can reflect a variety of underlying impairments including hemiparesis, hemiataxia, and loss of sensation. Similarly, the cognitive and behavioral outcome scores do not specify the nature of the corresponding cognitive and/or behavioral problems, complicating interpretation of impairment scores and associated lesion sites. Since this lack of granularity necessarily limits the specificity and interpretability of identified lesion-symptom relationships, future studies should prioritize the collection of more granular outcome measures.

Second, our sample may not have been sufficiently powered to allow for the identification of moderation effects, especially given the multiple comparisons corrections required by the voxel-wise analyses. Power for moderation effects is often substantially lower than for main effects^91,92^, and for voxel-level analyses, the power at any given voxel depends on the number of patients with damage to that voxel and the age distributions of patients with vs. without damage. Despite our large sample and broad lesion coverage, most voxels were damaged in fewer than 30 patients, complicating the estimation of stable voxel-level moderation effects. Even for ordinary multiple linear regressions, it is very difficult to accurately estimate power for moderation effects due to their dependence on multiple factors including the main effect magnitudes, the interaction effect magnitude, the correlation structure, and the shape of the interaction^92^, and this difficulty is amplified by our use of ordinal logistic regression models. It is therefore possible that our analyses lacked sufficient power at many or even most voxels, especially if age at stroke onset only weakly and partially modifies^92^ the effects of lesion location, as suggested by the conditional effects analyses for the hemisphere-level moderation effects (**Figure 2B**; **Supplementary Figure 3**).

Finally, despite our large patient sample, patients with perinatal/neonatal stroke were not included. While this was by design in that the parent study focused on childhood stroke, it complicates direct comparisons against other studies that have evaluated effects of age at stroke onset in mixed samples that include both perinatal/neonatal and childhood stroke survivors. We therefore emphasize that our conclusions cannot be generalized beyond childhood stroke. Evaluating whether similar results are obtained in a sample that includes both neonatal and childhood stroke is an important avenue for future work.

### Conclusions

We characterized how chronic neurological outcomes relate to age at stroke onset and lesion characteristics in childhood stroke survivors, and conducted the largest and most comprehensive lesion-symptom mapping study of childhood stroke to date. While younger age at stroke onset was consistently associated with worse outcomes when considered in isolation, multivariable models suggested this was partially explained by associations with lesion characteristics. We also found that the detrimental effect of extensive left hemisphere damage on expressive language outcomes increased with older age at stroke onset, and we identified subthreshold effects suggesting the possibility of interactions between age at onset and lesion characteristics for other outcome domains. Lastly, we identified robust fine-grained lesion-symptom associations with domain-level neurological outcomes, highlighting lesion location as a potentially important factor contributing to childhood stroke outcomes. Upon publication, we will also share the lesion-symptom maps generated by this study as a public resource for clinicians and researchers.

## Data availability

Data are available on request to the senior co-authors or via submitting an International Pediatric Stroke Study (IPSS) research proposal form, available at https://internationalpediatricstroke.org/ipss-research/.

Scripts that generated the results reported in the main text and supplement are available at: https://github.com/jcgriffis/pediatric_lesion_mapping. Statistical maps will be uploaded to NeuroVault after publication as a pre-print.

## Acknowledgments

We would like to thank Maya Dhar as the SCOUTS senior research coordinator, the research coordinators at all SCOUTS sites, and the patients and their families for participation in the VIPS and SCOUTS studies. We would also like to acknowledge Amanda Robertson as the IPSS Central Stroke Imaging Lab Manager and the Auxilium Foundation for IPSS Central support.

## Appendix

### Coinvestigators

| Name | Location | Role | Contribution |
|---|---|---|---|
| Clara Di Germiano | Vitalant Research Institute, San Francisco, CA | Co-Investigator | Laboratory Core |
| Philip J. Norris | Vitalant Research Institute, San Francisco, CA | Co-Investigator | Laboratory Core Lead |
| Adam L. Numis | UCSF, San Francisco, CA | Co-Investigator | Epileptologist |
| Lauren A. Beslow | Children’s Hospital of Philadelphia, Philadelphia, PA | Site Investigator | Enrollment and data collection |
| Paola Pergami | Children’s National Hospital, Washington DC | Site Investigator | Enrollment and data collection |
| Rachelle Herring | Cook Children’s Hospital, Fort Worth, TX | Site Investigator | Enrollment and data collection |
| Jenny Wilson | Oregon Health & Science University, Portland, OR | Site Investigator | Enrollment and data collection |
| Catherine Amlie-Lefond | Seattle Children’s Hospital, Seattle, WA | Site Investigator | Enrollment and data collection |
| Neil Friedman | Phoenix Children’s Hospital, Phoenix, AZ | Site Investigator | Enrollment and data collection |
| Marcie Files | Children’s Mercy Hospital, Kansas City, MO | Site Investigator | Enrollment and data collection |
| Beth Anne Cavanaugh | Le Bonheur Children’s Hospital, Memphis, TN | Site Investigator | Enrollment and data collection |
| Ryan Felling | Johns Hopkins Children’s Center, Baltimore, MD | Site Investigator | Enrollment and data collection |
| Lisa Sun | Johns Hopkins Children’s Center, Baltimore, MD | Site Investigator | Enrollment and data collection |
| Andrea Pardo | Lurie Children’s Hospital of Chicago, Chicago, IL | Site Investigator | Enrollment and data collection |

