## Supplementary Material for "Effects of age and lesion characteristics on chronic outcomes after childhood stroke: a large-scale study of 469 childhood stroke survivors"

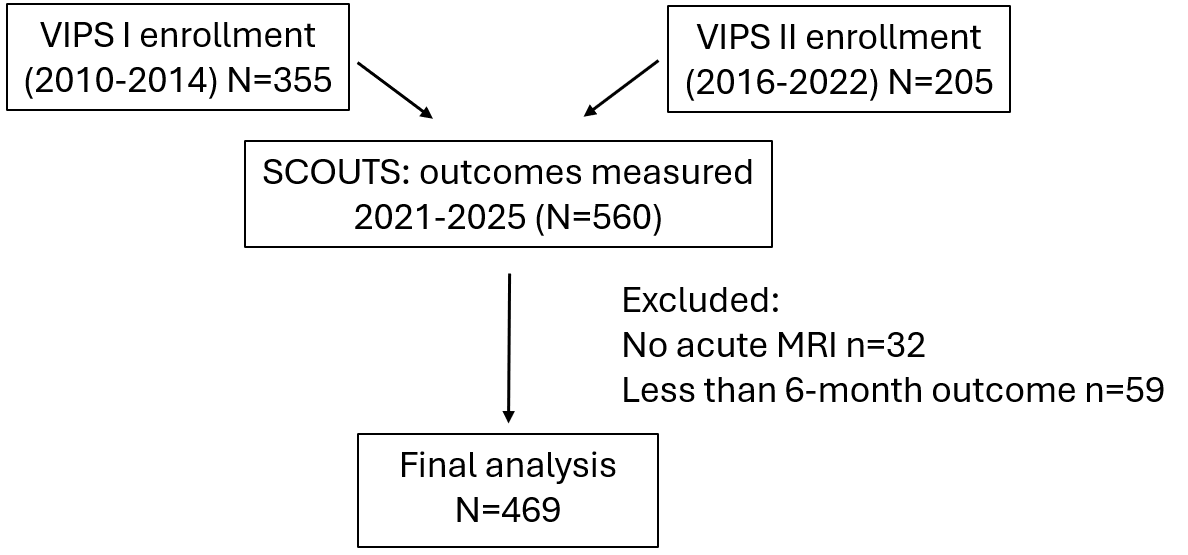

**Supplementary Figure 1.** Flowchart showing cohort breakdown, total data collected, exclusions, and patients included in the final analyses.

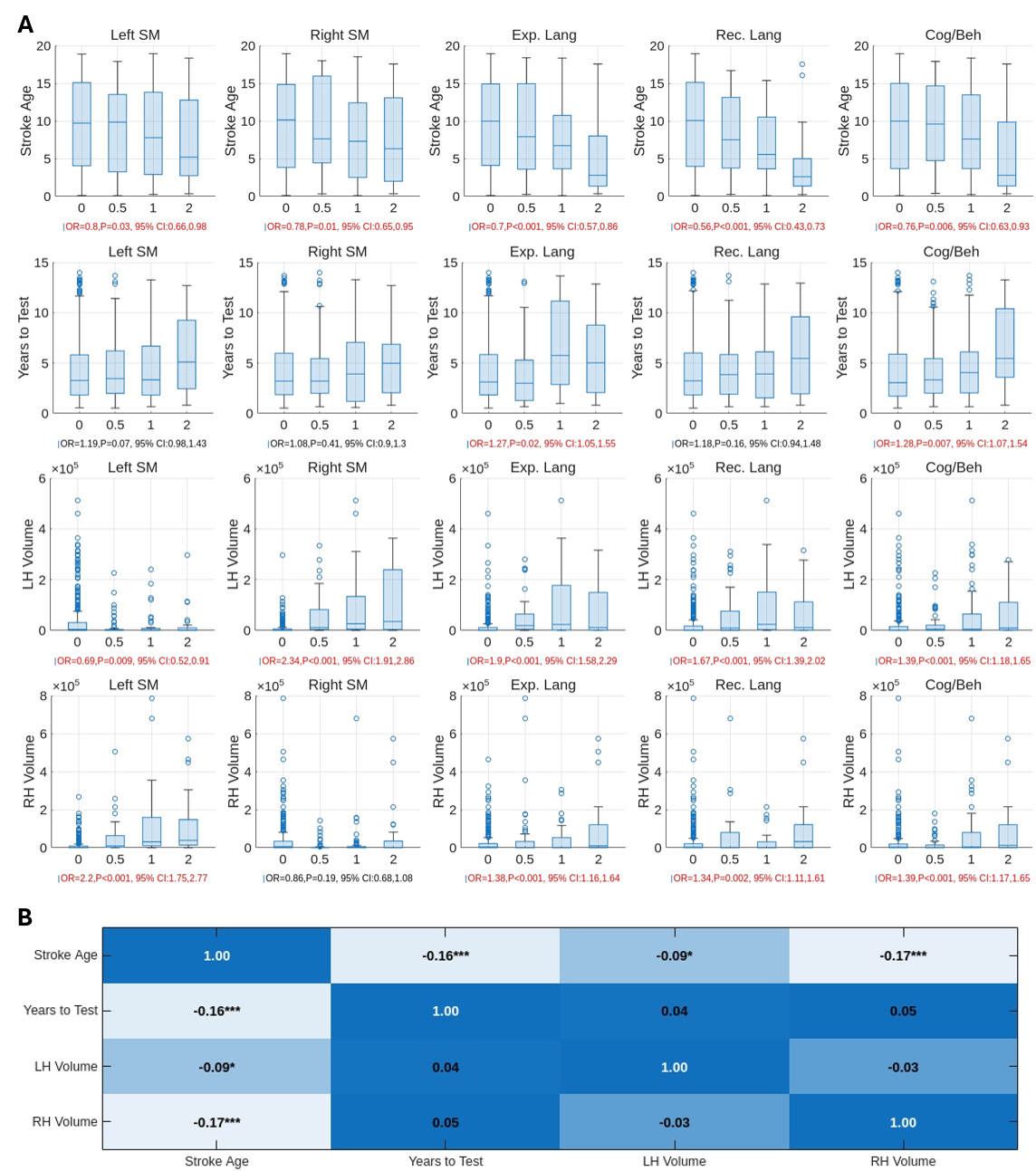

**Supplementary Figure 2.** Bivariate relationships among variables of interest. **A.** The boxplots show the stroke onset age distributions for each level of each outcome domain. Higher outcome scores correspond to more severe impairment. Cumulative Odds Ratios (ORs) along with per-comparison (i.e. uncorrected) *P*-values and 95% CIs obtained from single-variable ordinal logistic regression models of each outcome (columns) are shown for each predictor (rows) under the x-axis of each plot. Statistically significant results at a per-comparison *P*-value threshold of 0.05 are shown in red text, and non-significant results are shown in black text. **B.** Bivariate Pearson correlations are shown between each pair of predictors. Asterisks correspond to statistically significant correlations at a per-comparison threshold of *P*<0.05. **P*<0.05, ***P*<0.01, ****P*<0.001

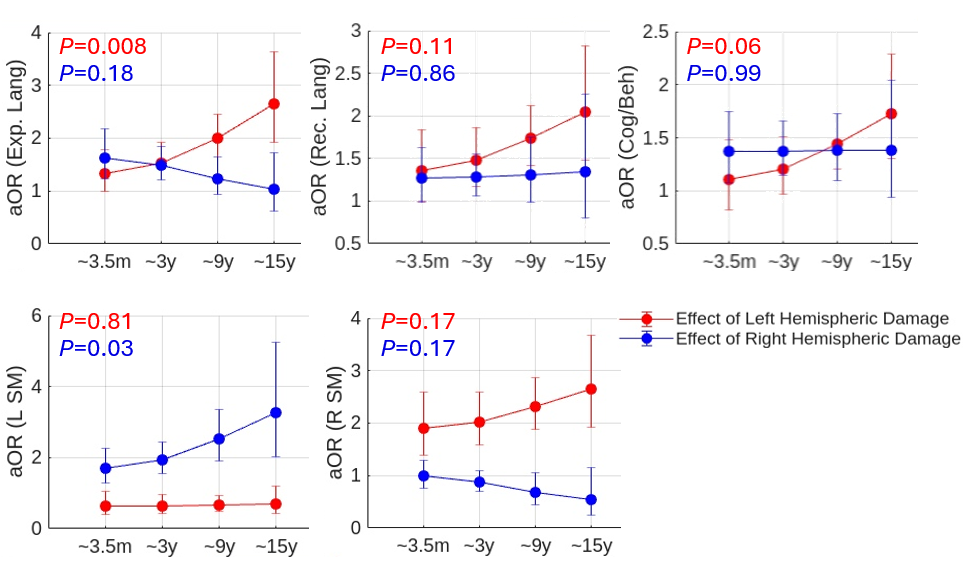

**Supplementary Figure 3.** Post-hoc conditional effects analyses for hemispheric lesion volumes at different levels of stroke onset age. Conditional effects analyses are shown for the age-by-hemispheric lesion volume interaction effects from Model 4 of each outcome. Adjusted cumulative odds ratios (aOR) and 95% CIs are shown for hemispheric lesion volume effects at age levels defined as -1.5, -1, 0, and 1 standard deviation units from the mean age at stroke onset, corresponding to ~3.5m, ~3y, ~9y, and ~15y stroke onset ages. The only statistically significant interaction effect in the main analyses was observed between age at stroke onset and left hemispheric lesion volume for expressive language in Model 3 (see Figure 2), but all putative interaction effects are visualized for completeness. *P*-values shown in the plot correspond to uncorrected *P*-values for the corresponding interaction terms in Model 4 of each outcome.

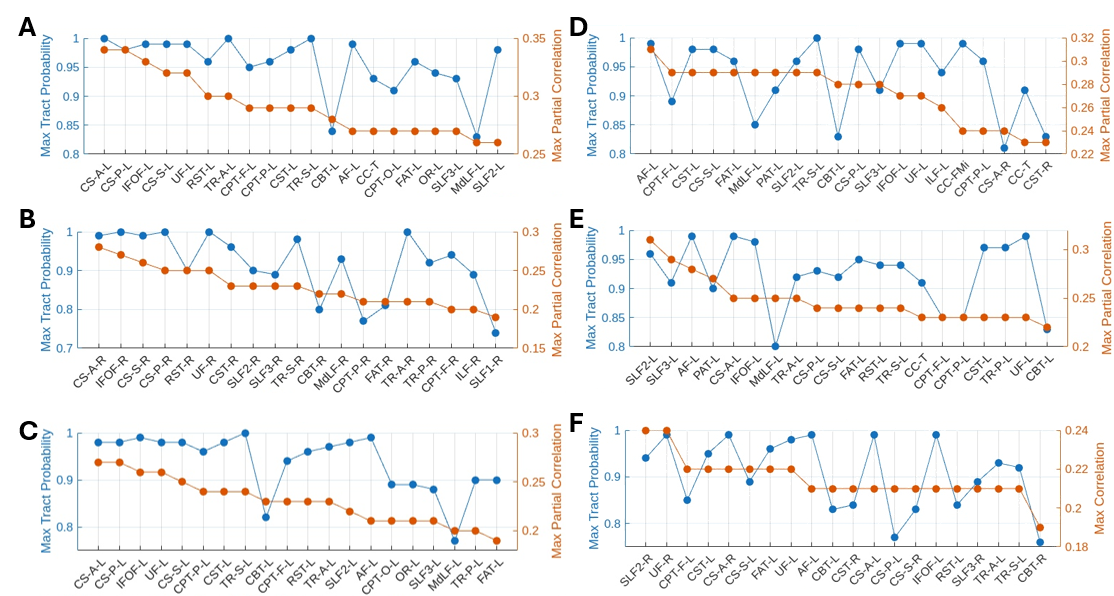

**Supplementary Figure 4.** Characterizing white matter results from the VLSM analyses. For each outcome, statistically significant voxels from the VLSM analyses were intersected with tract probability maps from the HCP-1065 tractography atlas (available at <https://brain.labsolver.org/hcp_trk_atlas.html>). Tracts were thresholded at 50% probability, and the maximum partial (or for Cognitive/Behavioral, full) correlation value was extracted along with the maximum tract probability value for intersecting voxels. For each outcome, plots show the maximum correlation values (orange) and the maximum tract probabilities (blue) for each of the top 20 tracts sorted by maximum correlation values. **A.** Contralateral sensorimotor. **B.** Left sensorimotor. **C.** Right sensorimotor. **D.** Expressive Language. **E.** Receptive Language. **F.** Cognitive/behavioral. Tract abbreviations are provided here (https://github.com/data-others/atlas/releases/download/hcp1065/abbreviation.xlsx).

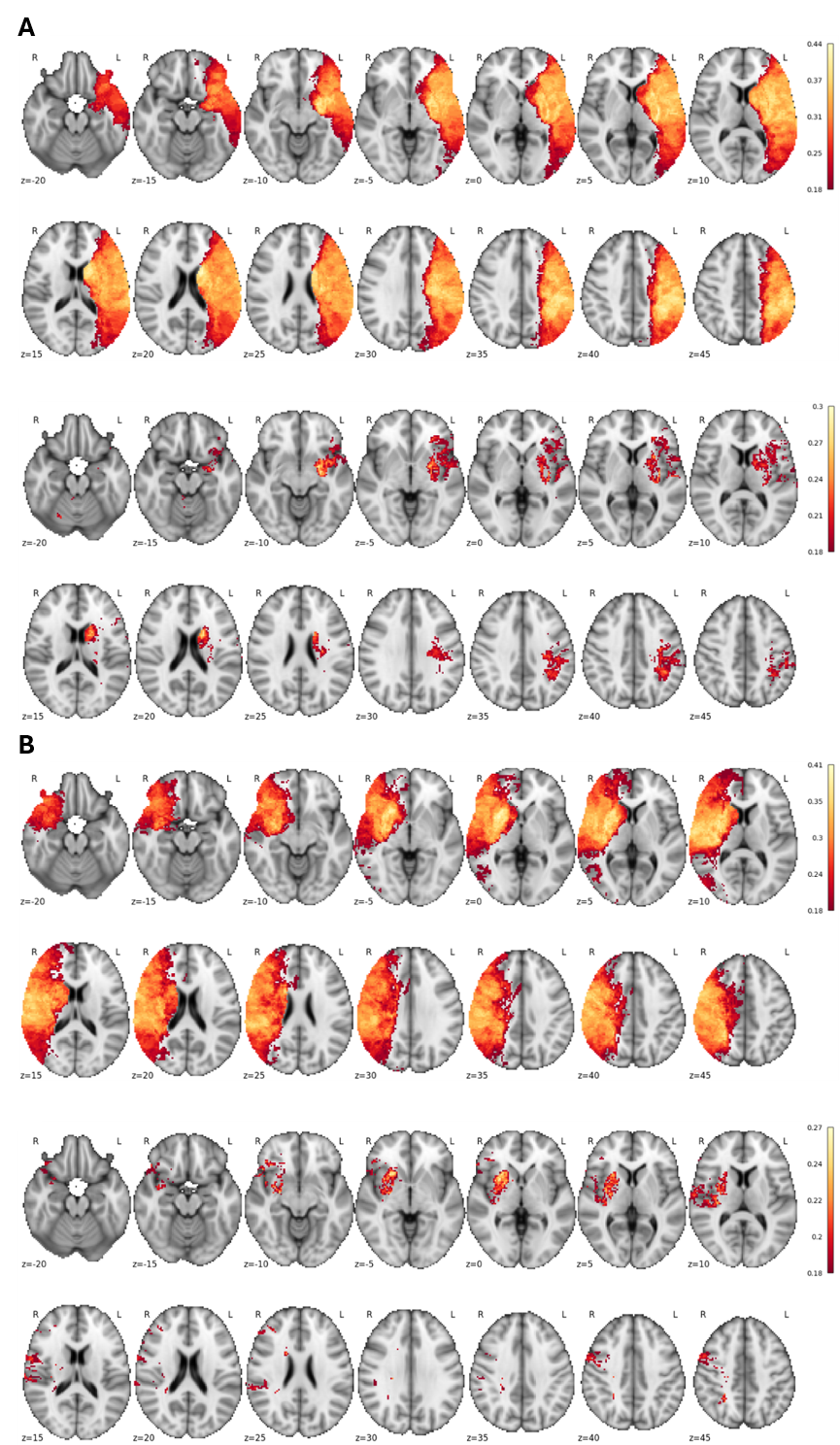

**Supplementary Figure 5.** Lesion-symptom mapping of left and right sensorimotor outcomes. **A.** Right sensorimotor outcome. Top panel - Full correlation results. Results are statistically significant at *P_FWE_* < 0.05 for *v*=100 voxels, with a post-hoc effective FDR of 0.002. Bottom panel - Partial correlation results. Results are statistically significant at *P_FWE_* < 0.05 for *v*=100 voxels, with a post-hoc effective FDR of 0.02. **B.** Left sensorimotor outcomes. Top panel - Full correlation results. Results are statistically significant at *P_FWE_*< 0.05 for *v*=100 voxels, with a post-hoc effective FDR of 0.002. Bottom panel - Partial correlation results. Results are statistically significant at *P_FWE_* < 0.05 for *v*=100 voxels, with a post-hoc effective FDR of 0.01.

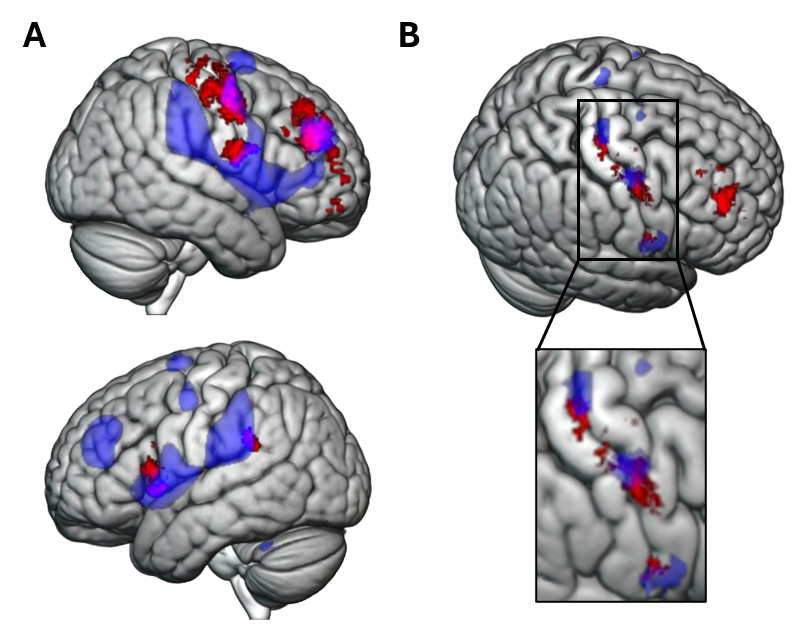

**Supplementary Figure 6.** Correspondence of cognitive and behavioral lesion-symptom mapping results with action mode and somato-cognitive action networks. **A.** The results for the cognitive and behavioral outcome are shown in red, and an action mode network (AMN) seed correlation map is overlaid in blue. The AMN seeds were located in the bilateral dACC and anterior insula, corresponding to the locations shown in Figure 1A from Dosenbach et al., (2025)^56^. Seed correlations were computed using the GSP-1000 dataset. The AMN seed correlation map was thresholded at *t* > 35.0 to isolate the core network. **B.** The cognitive and behavioral lesion-symptom map was thresholded at rho >= 0.20 to break up the larger pre-central gyrus/central sulcus clusters into smaller clusters around local peaks, shown in red. The inter-effector nodes (IENs) of the somato-cognitive action network (SCAN) are overlaid in blue. IEN nodes were obtained from the authors of Gordon et al., (2023)^57^.

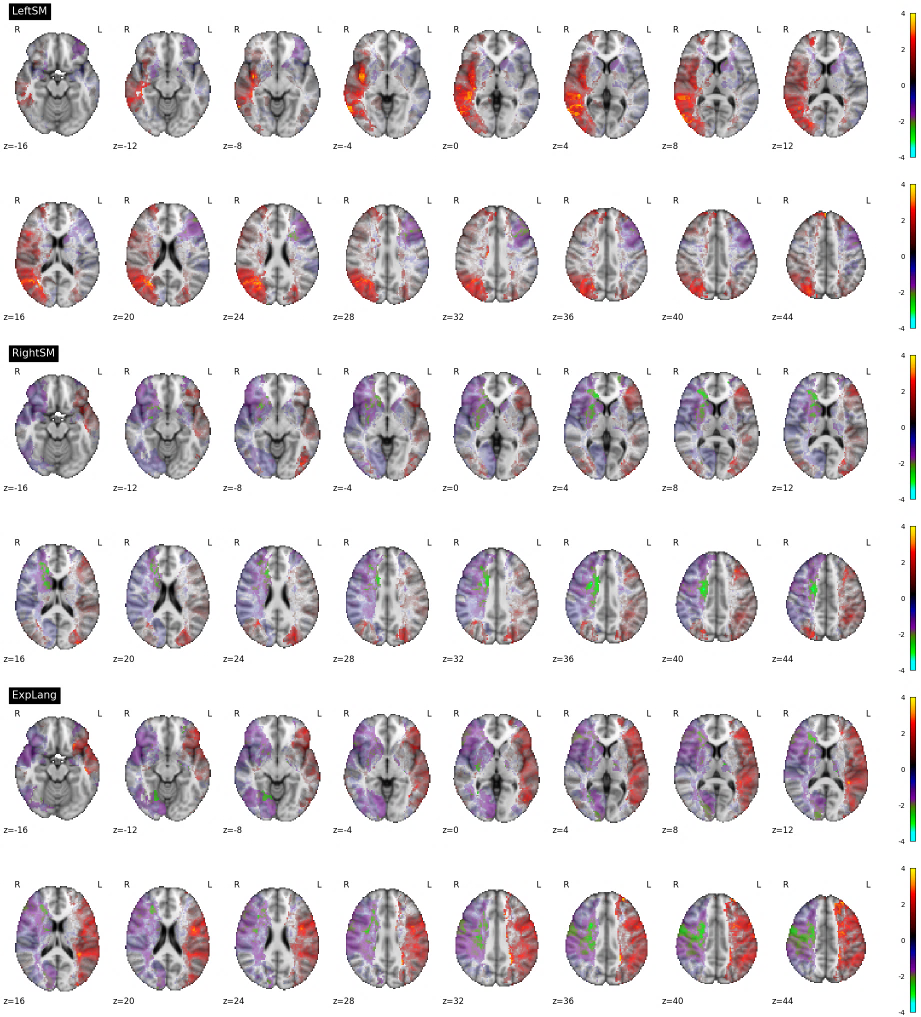

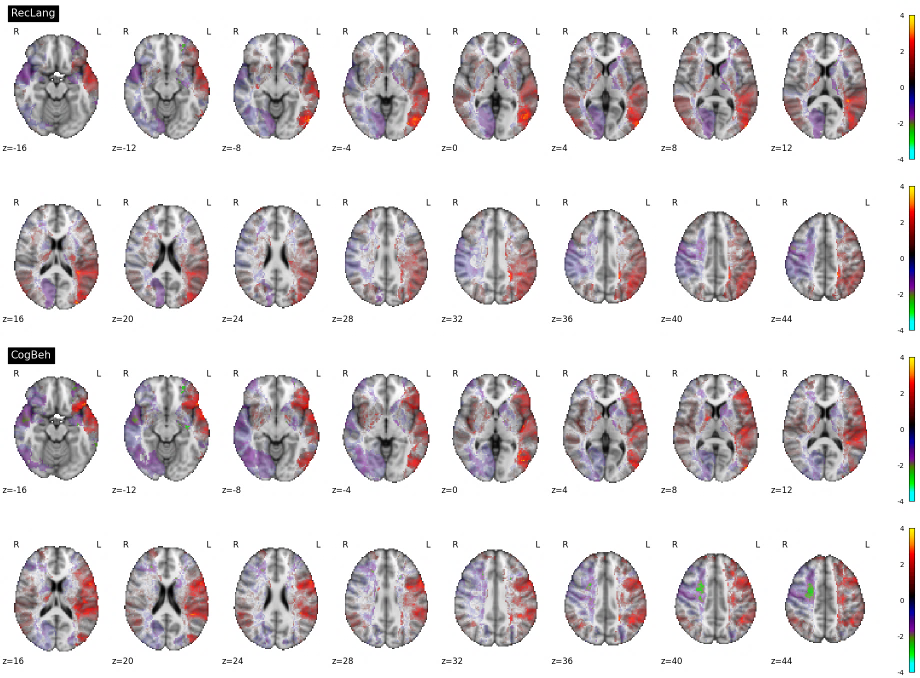

**Supplementary Figure 7.** Uncorrected statistical maps for voxel-wise moderation analyses. Maps show *t*-statistics for moderation effects from ordinal logistic regression models of left sensorimotor, right sensorimotor, expressive language, receptive language, and cognitive/behavioral outcomes. Maps are unthresholded, but voxels that did not survive an uncorrected threshold of *P*<0.01 (i.e. |*t|* < 2.58) are shown with variable transparency, becoming more opaque as they approach the threshold, while voxels surviving the threshold are rendered as opaque. Uncorrected p-value thresholds were determined using the parametric p-values obtained directly from the ordinal logistic regression models. Positive values suggest that the association between damage and chronic impairment increases with older age at stroke onset, all else held constant. Negative values suggest that the association between damage and chronic impairment decreases with increasing age at stroke onset, all else held constant.
